# Power and sample sizes estimation in clinical trials with treatment switching in intention-to-treat analysis: a simulation study

**DOI:** 10.1101/2022.09.07.22279686

**Authors:** Lejun Deng, Chih-Yuan Hsu, Yu Shyr

## Abstract

**Background:** Treatment switching, also called crossover, is common in clinical trials because of ethical concerns or other reasons. When it occurs and the primary objective is to identify treatment effects, the most widely used intention-to-treat analysis may lead to underpowered trials. Therefore, we plan to propose an approach to preview power reductions and to estimate sample sizes required for the desired power when treatment switching occurs in the intention-to-treat analysis.

**Methods:** We proposed a simulation-based approach and developed an R package to attain the purpose.

**Results:** We simulated a number of randomized trials incorporating treatment switching and investigated the impact of the relative effectiveness of the experimental treatment to the control, the switching probability, the allocation ratio, and the switching time on power reductions and sample sizes estimation. The switching probability and the switching time are key determinants for significant power decreasing and thus sample sizes surging to maintain the desired power. The sample sizes required in randomized trials absence of treatment switching varied from around three-fourths to one-twelfths of the sample sizes required in randomized trials allowing treatment switching as the probability of switched patients increased. The power reductions and sample sizes increase with the decrease of switching time.

**Conclusions:** The simulation-based approach not only provides a preview for power declining but also calculates the required sample size to achieve an expected power in the intention-to-treat analysis when treatment switching occurs. It will provide researchers and clinicians with useful information before randomized controlled trials are conducted.

## 1. Background

Clinical trials are often used to assess the effectiveness of new treatments. The most reliable clinical trial is the randomized controlled trials (RCT). In an RCT, patients are randomly assigned to the control group, where they receive a placebo or an existing standard treatment, or the experimental group, where they receive the experimental treatment. For treatments against diseases like cancer, the effectiveness of the new treatment is determined by comparing the observed survival time of patients in the experimental group to those in the control group. The power of a statistical test, which is the probability to correctly tell if the experimental treatment is better when it is truly better, is extremely important. Being able to correctly determine the effectiveness of an experimental treatment gives justification for its production, allows for better utilization of resources, and helps those who will benefit from the experimental treatment.

A common method used to interpret the data from a RCT is the intention-to-treat analysis (ITT). The ITT analysis includes all patients with randomization in statistical analysis and compares their responses to determine the effectiveness of the experimental treatment according to the treatment group that was initially assigned to them, regardless of what treatment they actually received. Patients, however, do not always comply with the treatment they were assigned. Because of ethical concerns or other reasons, they may switch treatments from the control group to the experimental group. For example, it may occur when a disease progresses or when the healthcare provider believes the patient’s prognosis will improve with the experimental treatment. When treatment switching, also called crossover, is permitted, the ITT analysis is confounded, which decreases the power of a statistical test. A simple and alternative approach, such as the per-protocol analysis, excludes the subjects who switch treatments from the statistical analysis. Nevertheless, this may heavily bias the analysis results if the subjects included and excluded in the analysis significantly differ in prognosis, i.e., treatment switching is associated with prognostic variables [1].

Various statistical methods dealing with treatment switching have been proposed. Law and Kaldor [2] proposed the adjusted Cox model by splitting the study population into four groups according to their initial and final treatment groups and by assuming different hazard functions with time dependent covariates defined by the time of switching treatment for the four group. Loeys and Goetghebeur [3] proposed the causal proportional hazards estimator by assuming that patients in one arm complete their treatment while the patients in the other arm either completely fulfills their treatment or does not fulfill their treatment at all. Robins and Tsiatis [4] proposed correcting for non-compliance by using the rank preserving structural accelerated failure time models (RPSFT). Based on the RPSFT, Branson and Whitehead [5] and Zhang and Chen [6] provided iterative parametric estimation and modified iterative parametric estimation methods, respectively, for fast and reliably estimating the treatment effect. Morden et al. [1] and Latimer et al. [7, 8] performed simulations studies to compare several adjustment methods mentioned above.

However, sample sizes estimation in most RCTs is still based on the assumption of no treatment switching and predetermined statistical tests, such as the logrank test in ITT analysis. Given the sample sizes estimated from no-switching designs, the ITT analysis may underestimate the positive treatment effect when treatment switching occurs. In this study, we proposed a simulation-based approach, *PowerSwitchingTrial*, to preview power reductions and calculate sample sizes required in RCTs allowing treatment switching. We investigated the impact of the relative effectiveness of the experimental treatment to the control, the switching probability, the allocation ratio, and the switching time on power reductions and sample sizes estimation. We found the switching probability and the switching time are key determinants for power decreasing and thus sample sizes increasing to maintain the desired power in ITT analysis. *PowerSwitchingTrial* is freely available at https://github.com/darwin-hub/PowerSwitchingTrial.

## 2. Methods

### 2.1 *PowerSwtichingTrial* overview

*PowerSwitchingTrial* is a simulation-based R package, developed for exploring statistical powers and sample sizes required in RCT trials allowing treatment switching when the logrank test is used in ITT analysis. The flowchart of *PowerSwitchingTrial* is shown in Fig. 1. *PowerSwtichingTrial* simulates a two-arm randomized clinical trial study with an accrual time of *T*_*a*_ and an additional follow-up time of *T*_*e*_ − *T*_*a*_, where *T*_*e*_ is the time from the start to the end of the trial. Participants are assumed to enter the study uniformly during the accrual time period, and soon be randomly assigned to one of the two treatment groups, the control group (*TX*_1_) or the experimental group (*TX*_2_), with an allocation ratio of *r* (*r* = *TX*_2_/*TX*_1_). The survival time of subjects in the control and experimental groups, denoted by *T*_1_ and *T*_2_, is assumed to follow exponential distributions with the median value of *m*_1_ and *m*_2_, respectively. The censoring time of subjects in the two groups, denoted by *C*_1_ and *C*_2_, is defined as the time from randomization until the end of the trial if the subjects do not experience the event of interest.

**Figure 1.**
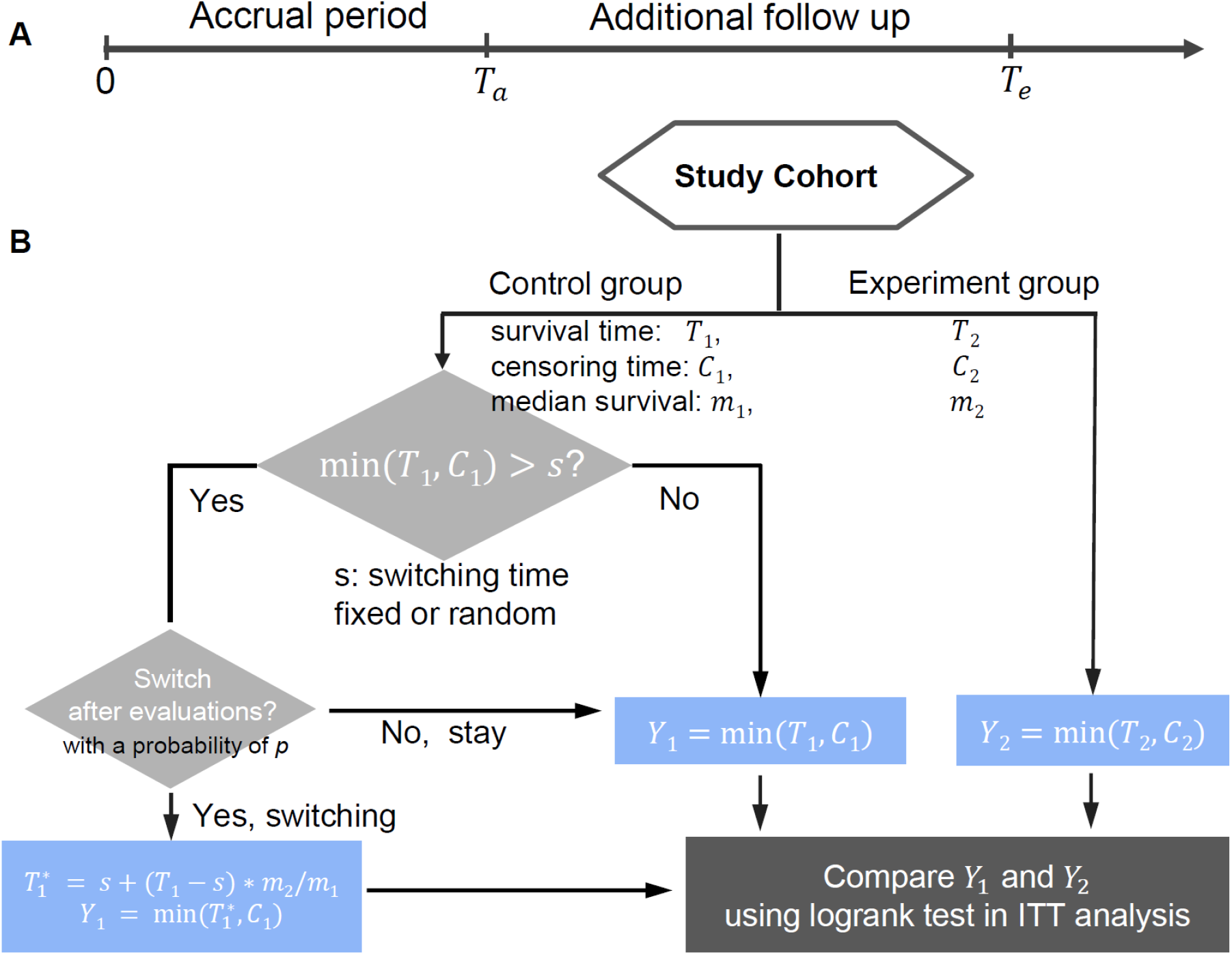
A flowchart of *PowerSwtichingTrial*.

*PowerSwtichingTrial* allows subjects in the control group have a chance to switch to the experimental group if predetermined conditions are satisfied. For example, if patients with cancer are observed to have a disease progression before death (assume death is the event of interest), they may switch from the standard treatment to the new treatment after disease progression. The switching time, denoted by *s*, is defined as the time from randomization until treatment switching. *s* is fixed or random, which depends on a practical application. For example, *s* will be fixed if there is a time set in a clinical trial to switch treatments, while s will be random if the switching time depends on disease progression. In the setting of random *s*, we assume *s* = *X* × *T*_1_, where *X* is a random variable generated from a beta distribution with the parameters of *a* and *b*, namely Beta(*a,b*). The two parameters control the relationship between *s* and *T*_1_. Moreover, *PowerSwtichingTrial* considers a parameter of *p* (switching probability) to define the probability that a subject who qualifies for treatment switching will switch from *TX*_1_ to *TX*_2_ after evaluations from physicians or health care professionals. The survival time for subjects starting from switching is assumed to increase by a constant multiplier of *A*, based on the rank preserving structural failure time model [4]. That is, the survival time of the subjects with treatment switching will be 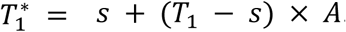. Therefore, the observable survival time *Y*_1_ for the subjects who are originally assigned to *TX*_1_ will be equal to *min*(*T*_1_, *C*_1_) without treatment switching, and 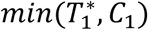 with treatment switching. The observable survival time *Y*_2_ for the subjects in *TX*_2_ will be equal to *min*(*T*_2_, *C*_2_). Finally, the logrank test is used to compare *Y*_1_ and *Y*_2_.

### 2.2 Two main functions of *PowerSwtichingTrial*

*PowerSwtichingTrial* provides two main functions, *LogRankTestMix2PowerMedian* and *LogRankTestMix2Nmedian. LogRankTestMix2PowerMedian* calculates the power, given sample sizes, the difference in the survival time between two groups, and the other parameters. *LogRankTestMix2NMedian* estimates sample sizes, given an expected power to detect the difference in the survival time between two groups, and the other required parameters.

*LogRankTestMix2PowerMedian*, a function to calculate the power, includes 13 input parameters to simulate a number of scenarios, *n, m*_1_, *m*_2_, *r, T*_*a*_, *T*_*e*_, *p, alpha, random, s, a, b*, and *reps. n* is the number of participants in the control group, *m*_1_, *m*_2_, *r, T*_*a*_, *T*_*e*_, and the switching probability (*p*) were defined in Section 2.1, *alpha* is the significance level, and *reps* is the number of computer simulations. The *random* is a logic indicator, FALSE or TRUE. If *random* = FALSE, the switching time is fixed and *s* is required; if *random* = TRUE, the switching time is random and the parameters of *a* and *b* in the beta distribution are required. The function will return the power, that is, the average number of rejecting the null hypothesis which suggests no difference in survival time between the two treatment groups across *reps* simulations.

*LogRankTestMix2Nmedian is* a function to calculate the sample sizes. *LogRankTestMix2NMedian* uses the same parameters as *LogRankTestMix2PowerMedian*, except the parameter of *n* that is replaced by *power, lower*, and *upper*. Lower and upper is the minimum and maximum sample sizes that users set to explore. The function uses the bisection method to find the sample size (*n*) required between the initial *lower* and *upper* bounds.

## 3. Results

A number of scenarios were generated to explore the effect of treatment switching on powers and sample sizes estimation in ITT analysis, which included the relative different effectiveness of the experimental treatment to the control, the different switching probability, the different allocation ratio, and the fixed/random switching time. In each scenario, the accrual time *T*_*a*_ was set to 3, the study time *T*_*e*_ was set to 5, and *m*_1_ was set to 1. Under no treatment switching, the expected power to detect the survival difference between two treatment groups was set as 0.8 and the significance level was set as 0.05. The number of computer simulations was set at 5,000.

### 3.1 Effect of the relative effectiveness on power and sample sizes estimation

We first investigated the impact of the relative effectiveness of the experimental treatment to the control (*m*_2_/*m*_1_) on power and sample sizes estimation, which ranged at 1.25, 1.5, 1.75, and 2 (Supplementary Tables s1-s3). We found the power decreased and the ratio of sample sizes with treatment switching to those absence of switching (*n*_*s*_/*n*_*no*−*s*_) increased slightly with the rising relative effectiveness (Fig. 2). For example, given *r* = 1, *p* = 0.4, and *s* = 0.5, the ratio increased from 1.94 to 2.07 and the power decreased from 0.52 to 0.50 when the relative effectiveness grew from 1.25 to 2 (Supplementary Table s1).

**Figure 2.**
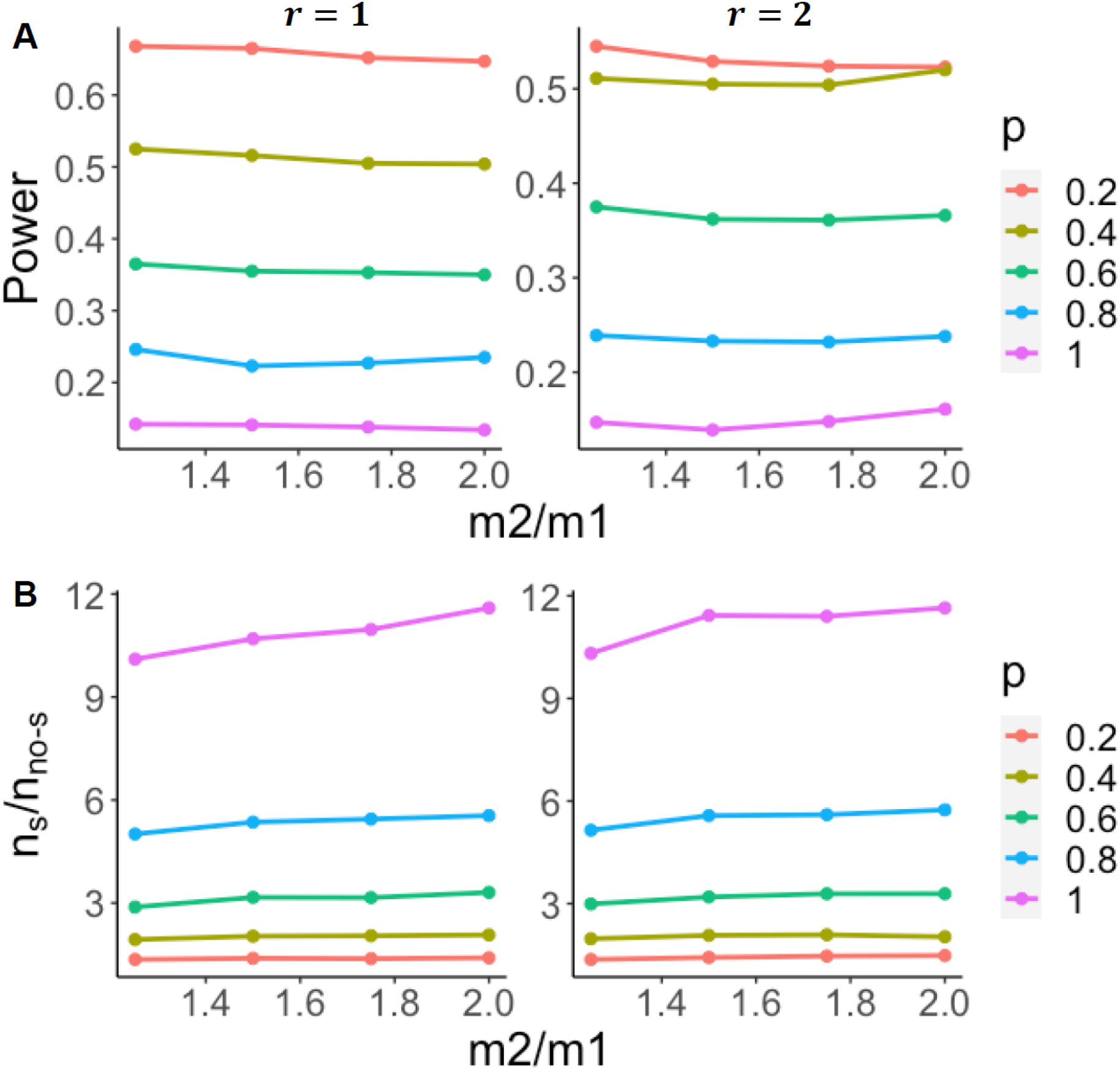
Effect of relative effectiveness on power (A) and sample sizes estimation (B). *s* = 0.5.

### 3.2 Effect of the switching probability on power and sample sizes estimation

We further evaluated the effect the switching probability (*p*) on power and sample sizes estimation, which was set at 0.2, 0.4, 0.6, 0.8, and 1.0. We found the power decreased and the ratio of sample sizes (*n*_*s*_/*n*_*no*−*s*_) increased significantly with the enlarging switching probability (Fig. 3). For example, the ratio of sample sizes increased from ∼1.4 to ∼11.6 and the power decreased from ∼0.7 to ∼0.1 When the switching probability increased from 0.2 to 1.0 (Supplementary Table s1). The result indicates that the switching probability is critical for power and sample sizes estimation. A small change of the switching probability would require much more sample sizes to maintain the expected power.

**Figure 3.**
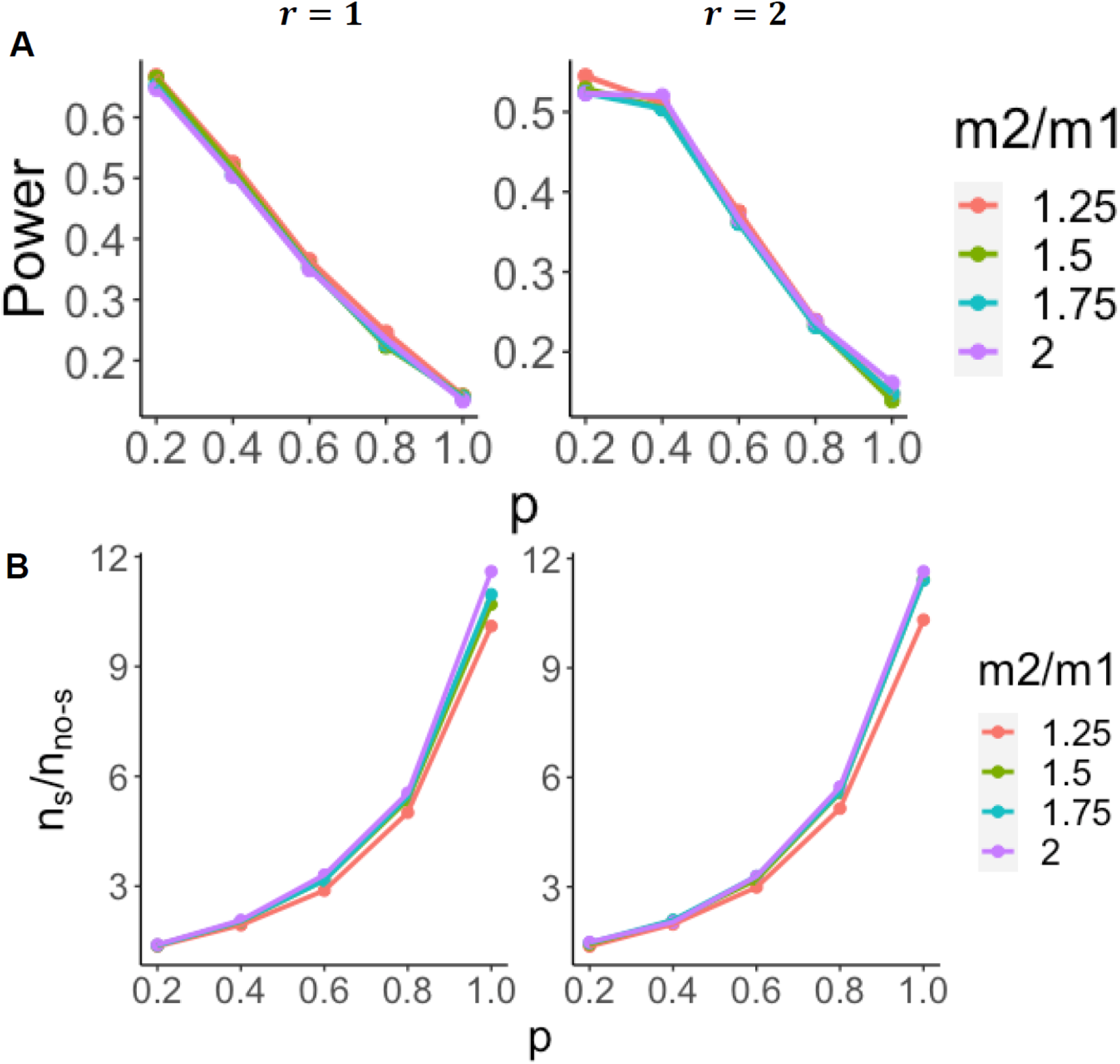
Effect of switching probability on power (A) and sample sizes estimation (B). *s* = 0.5.

### 3.3 Effect of the allocation ratio on power and sample sizes estimation

Moreover, we examined the effect of the allocation ratio (*r*) on power and sample sizes estimation, which was set at 1 and 2. The sample size required for the control group decreased as the allocation ratio increased from 1 to 2, the total sample size increased though. The ratio of sample sizes (*n*_*s*_/*n*_*no*−*s*_) increased slightly but the power didn’t show a clear increasing or decreasing pattern. For example, under *s* = 0.5, given *p* = 0.2, the power with *r* = 1 was larger than that with *r* = 2. In contrast, the power with *r* = 1 was smaller than that with *r* = 2 given *p* = 1 (Supplementary Table s1).

### 3.4 Effect of the switching time on power and sample sizes estimation

Finally, we explored the effect of the switching time on power and sample sizes estimation. The switching time (*s*) was either set at a fixed value or generated by random. The switching time was set at 0.5 and 1 if fixed, otherwise the switching time was generated by a product of survival time of the control group and a random variable from a beta distribution with shape parameters of *a* = 2 and *b* = 2. The mean of the random switching time was 0.5. Overall, the ratios of sample sizes (*n*_*s*_/*n*_*no*−*s*_) using random switching time were smaller than those using the fixed switching time of 0.5 (Fig. 4 and Supplementary Tables s1 and s2). The ratio of sample sizes (*n*_*s*_/*n*_*no*−*s*_) increased and power decreased significantly when the fixed switching time moved to an earlier time point (from 1 to 0.5), especially when the switching probability *p* is greater than 0.6 (Fig.4 and Supplementary Table s1 and s3). For example, the ratio of sample sizes decreased from ∼11 to ∼3 when the switching time moved from 0.5 to 1 under *p* = 1, *r* = 1, and *m*2/*m*1 = 1.5 (Fig. 4). The results indicated that the switching time is also a key factor for power reductions and sample sizes increase in clinical trials with treatment switching.

**Figure 4.**
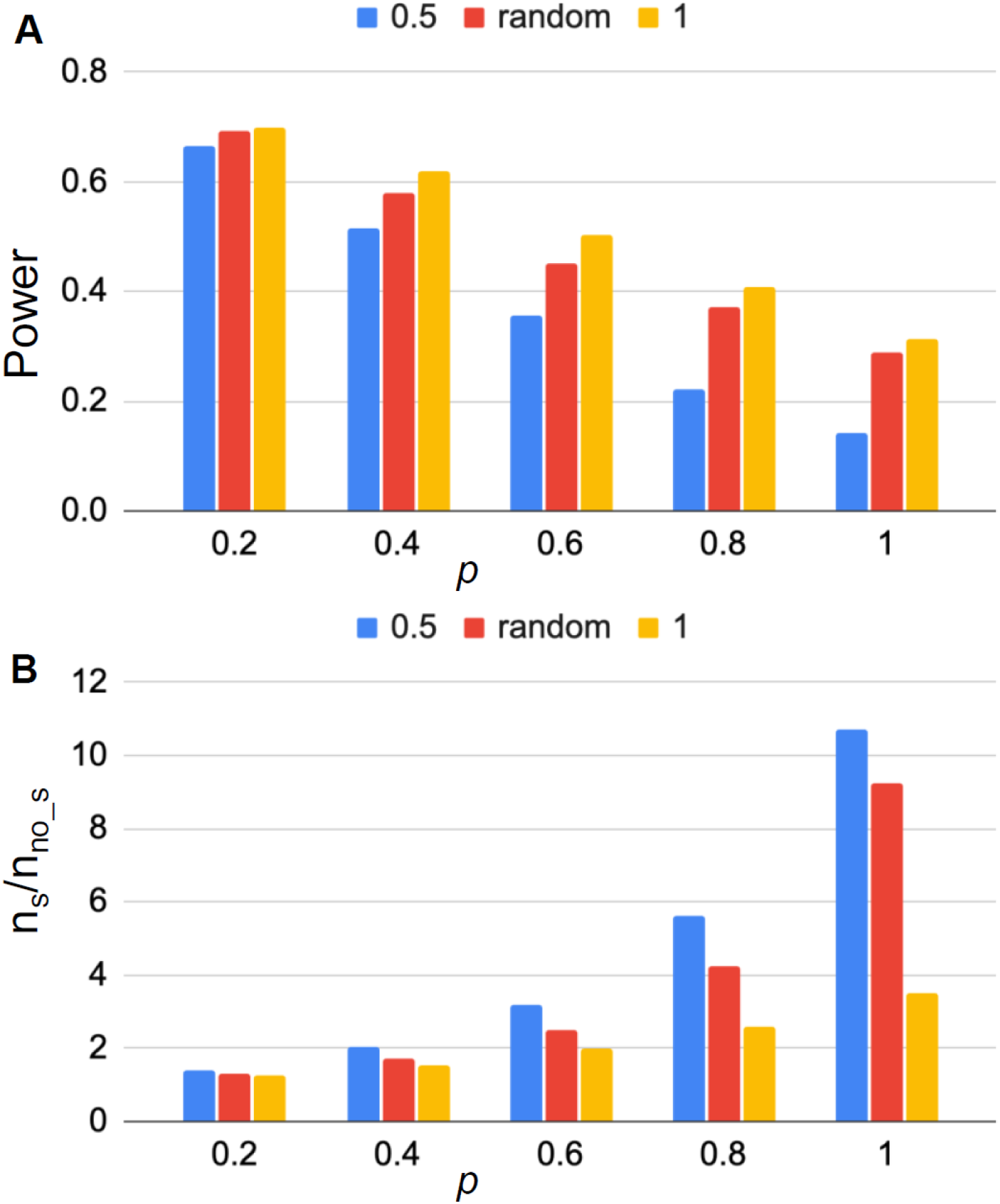
Effect of switching time on power (A) and sample sizes estimation (B). *s* = 0.5 (blue), *s* = 1 (orange) or s is random based on a beta distribution with mean value of 0.5 (red) under *r* = 1 and *m*2/*m*1 = 1.5.

## 4. Discussion

Our simulation study showed a standard ITT analysis failing to consider treatment switching resulted in a significant reduction of statistical powers especially when the switching probability is high and the switching time is early, which indicated that a much larger sample size was required to maintain the expected power. The sample sizes required in RCTs absence of treatment switching varied from around three-fourths to one-twelfths of the sample sizes required in RCTs allowing treatment switching as the probability of switched patients increased. The growth of the relative effectiveness and the switching probability increased the ratio of samples and decreased the power, while the later switching time decreased the ratio of sample sizes and increased the power. The allocation ratio was slightly associated with the increasing sample sizes but was not associated with the decreasing power.

*PowerSwitchingTrial* assumed the effects of the experimental treatment were the same for the subjects switched from *TX*_1_ to *TX*_2_ and those initially assigned to *TX*_2_, which is the same as the “common treatment effect” assumption made by RPSFTM [8]. In some cases, the assumption may be problematic. The subjects switched from *TX*_1_ to *TX*_2_ may be associated with worse survival. Properly adjusting the accelerated factor *A* may be useful to fit the scenario. Multiplying a constant that is less than 1 to *A* may be a solution, but it is difficult to determine the constant value before clinical trials even if we can borrow the information from previous similar studies.

The assumption of exponential distributions for survival time also limits the flexibility and practice of *PowerSwitchingTrial*. Survival times do not always follow exponential distributions well. Weibull or gamma distribution may be alternatives to exponential distributions. However, it is not easy for clinicians to determine a non-exponential distribution with at least two parameters. Median survival or mean survival is still commonly used, which leads to favoring of exponential distributions. In addition, *PowerSwitchingTrial* assumed the censoring time depended only on the entry time and study time although other factors that would cause censoring include a small proportion of patients dropping out from the study and a loss of follow up. This was due to no need for additionally assuming a probability distribution for the censoring time.

The current version of *PowerSwitchingTrial* is built for two-arm RCTs. It can be extended to multi-arm RCTs, straightforwardly but not simply. More parameters need to be determined in multi-arm clinical trial designs. We plan to extend *PowerSwitchingTrial* for multi-arm RCTs in the near future.

## 5. Conclusions

In this study, we proposed a simulation-based approach, *PowerSwitchingTrial*, to evaluate statistical powers and sample sizes required in RCTs allowing switching in ITT analysis when the logrank test is used. The approach not only provides a preview for power declining but also calculates the required sample size to achieve an expected power when treatment switching occurs. It will provide researchers and clinicians with useful information before RCTs are conducted.

## Data Availability

none

## Abbreviations

ITT: intention-to-treat
RCT: randomized controlled trials
RPSFT: rank preserving structural accelerated failure time models

## Declarations

### Ethics approval and consent to participate

Not applicable.

### Consent for publication

Not applicable.

### Availability of data and materials

Additional file 1: Supplementary Tables s1-s3.

### Authors’ contributions

LD and CYH were major contributors in writing the manuscript and developing the R package. YS was the major contributor in the conception and design of the work.

## Acknowledgements

Not applicable.

