## Supplemental Table s1-s3 for "Power and sample sizes estimation in clinical trials with treatment switching in intention-to-treat analysis: a simulation study"

**Table s1:** Simulation results for powers and sample sizes under $s = 0.5$.

**Table s2.** Simulation results for powers and sample sizes under $s = X \times T_1$, where $X \sim Beta(a, b)$, $a = 2$ and $b = 2$.

**Table s3:** Simulation results for powers and sample sizes under $s = 1$.

**Table s1.** Simulation results for powers and sample sizes under $s = 0.5$.

| $r$ | $p$ | $m_2/m_1$ | $n_{no-s}$ (no switch) | $n_S$ (switch) | Ratio of $n_s/n_{no-s}$ | Power based on $n_{no-s}$ |
|---|---|---|---|---|---|---|
| 1 | 0.2 | 1.25 | 369 | 501 | 1.358 | 0.668 |
| | | 1.50 | 115 | 160 | 1.391 | 0.665 |
| | | 1.75 | 63 | 87 | 1.396 | 0.652 |
| | | 2.00 | 42 | 59 | 1.404 | 0.647 |
| 1 | 0.4 | 1.25 | 369 | 715 | 1.938 | 0.525 |
| | | 1.50 | 115 | 234 | 2.035 | 0.516 |
| | | 1.75 | 63 | 129 | 2.048 | 0.505 |
| | | 2.00 | 42 | 87 | 2.071 | 0.504 |
| 1 | 0.6 | 1.25 | 369 | 1064 | 2.883 | 0.365 |
| | | 1.50 | 115 | 364 | 3.165 | 0.355 |
| | | 1.75 | 63 | 199 | 3.159 | 0.353 |
| | | 2.00 | 42 | 139 | 3.310 | 0.35 |
| 1 | 0.8 | 1.25 | 369 | 1849 | 5.011 | 0.246 |
| | | 1.50 | 115 | 616 | 5.617 | 0.223 |
| | | 1.75 | 63 | 343 | 5.444 | 0.227 |
| | | 2.00 | 42 | 233 | 5.548 | 0.235 |
| 1 | 1.0 | 1.25 | 369 | 3728 | 10.103 | 0.142 |
| | | 1.50 | 115 | 1230 | 10.696 | 0.141 |
| | | 1.75 | 63 | 691 | 10.968 | 0.138 |
| | | 2.00 | 42 | 487 | 11.595 | 0.134 |
| 2 | 0.2 | 1.25 | 271 | 371 | 1.369 | 0.545 |
| | | 1.50 | 82 | 117 | 1.427 | 0.529 |
| | | 1.75 | 45 | 66 | 1.467 | 0.524 |
| | | 2.00 | 31 | 46 | 1.484 | 0.523 |
| 2 | 0.4 | 1.25 | 271 | 535 | 1.974 | 0.511 |
| | | 1.50 | 82 | 170 | 2.073 | 0.505 |
| | | 1.75 | 45 | 94 | 2.089 | 0.504 |
| | | 2.00 | 31 | 63 | 2.032 | 0.520 |
| 2 | 0.6 | 1.25 | 271 | 810 | 2.980 | 0.375 |
| | | 1.50 | 82 | 262 | 3.195 | 0.362 |
| | | 1.75 | 45 | 148 | 3.289 | 0.361 |
| | | 2.00 | 31 | 102 | 3.290 | 0.366 |
| 2 | 0.8 | 1.25 | 271 | 1395 | 5.148 | 0.239 |
| | | 1.50 | 82 | 457 | 5.573 | 0.233 |
| | | 1.75 | 45 | 252 | 5.600 | 0.232 |
| | | 2.00 | 31 | 178 | 5.742 | 0.238 |

| | | | | | | |
|---|---|---|---|---|---|---|
| 2 | 1.0 | 1.25 | 271 | 2796 | 10.314 | 0.147 |
| | | 1.50 | 82 | 937 | 11.427 | 0.139 |
| | | 1.75 | 45 | 513 | 11.400 | 0.148 |
| | | 2.00 | 31 | 361 | 11.645 | 0.161 |

**Table s2.** Simulation results for powers and sample sizes under $s = X \times T_1$, where $X \sim Beta(a,b)$, $a = 2$ and $b = 2$.

| $r$ | $p$ | $m_2/m_1$ | $n_{no-s}$ (no switch) | $n_S$ (switch) | Ratio of $n_s/n_{no-s}$ | Power based on $n_{no-s}$ |
|---|---|---|---|---|---|---|
| 1 | 0.2 | 1.25 | 369 | 455 | 1.233 | 0.702 |
| | | 1.50 | 115 | 150 | 1.304 | 0.694 |
| | | 1.75 | 63 | 83 | 1.317 | 0.686 |
| | | 2.00 | 42 | 56 | 1.333 | 0.676 |
| 1 | 0.4 | 1.25 | 369 | 611 | 1.543 | 0.598 |
| | | 1.50 | 115 | 200 | 1.739 | 0.578 |
| | | 1.75 | 63 | 111 | 1.762 | 0.592 |
| | | 2.00 | 42 | 77 | 1.833 | 0.567 |
| 1 | 0.6 | 1.25 | 369 | 863 | 2.339 | 0.484 |
| | | 1.50 | 115 | 288 | 2.504 | 0.450 |
| | | 1.75 | 63 | 163 | 2.587 | 0.452 |
| | | 2.00 | 42 | 113 | 2.690 | 0.432 |
| 1 | 0.8 | 1.25 | 369 | 1438 | 3.897 | 0.377 |
| | | 1.50 | 115 | 489 | 4.252 | 0.371 |
| | | 1.75 | 63 | 282 | 4.476 | 0.363 |
| | | 2.00 | 42 | 207 | 4.929 | 0.338 |
| 1 | 1.0 | 1.25 | 369 | 3051 | 8.268 | 0.299 |
| | | 1.50 | 115 | 1061 | 9.226 | 0.289 |
| | | 1.75 | 63 | 653 | 10.365 | 0.270 |
| | | 2.00 | 42 | 488 | 11.619 | 0.253 |
| 2 | 0.2 | 1.25 | 271 | 342 | 1.262 | 0.695 |
| | | 1.50 | 82 | 107 | 1.305 | 0.693 |
| | | 1.75 | 45 | 59 | 1.311 | 0.688 |
| | | 2.00 | 31 | 41 | 1.323 | 0.687 |
| 2 | 0.4 | 1.25 | 271 | 458 | 1.690 | 0.604 |
| | | 1.50 | 82 | 148 | 1.805 | 0.589 |
| | | 1.75 | 45 | 82 | 1.822 | 0.589 |
| | | 2.00 | 31 | 57 | 1.839 | 0.564 |
| 2 | 0.6 | 1.25 | 271 | 660 | 2.435 | 0.475 |
| | | 1.50 | 82 | 211 | 2.695 | 0.472 |
| | | 1.75 | 45 | 121 | 2.689 | 0.444 |
| | | 2.00 | 31 | 85 | 2.742 | 0.436 |
| 2 | 0.8 | 1.25 | 271 | 1053 | 3.886 | 0.382 |
| | | 1.50 | 82 | 350 | 4.268 | 0.365 |
| | | 1.75 | 45 | 208 | 4.622 | 0.357 |

| | | | | | | |
|---|---|---|---|---|---|---|
| | | 2.00 | 31 | 150 | 4.839 | 0.330 |
| 2 | 1.0 | 1.25 | 271 | 2267 | 8.365 | 0.311 |
| | | 1.50 | 82 | 803 | 9.793 | 0.277 |
| | | 1.75 | 45 | 469 | 10.422 | 0.275 |
| | | 2.00 | 31 | 375 | 12.097 | 0.273 |

**Table s3.** Simulation results for powers and sample sizes under $s = 1$.

| $r$ | $p$ | $m_2/m_1$ | $n_{no-s}$ (no switch) | $n_S$ (switch) | Ratio of $n_s/n_{no-s}$ | Power based on $n_{no-s}$ |
|---|---|---|---|---|---|---|
| 1 | 0.2 | 1.25 | 369 | 446 | 1.209 | 0.716 |
| | | 1.50 | 115 | 143 | 1.243 | 0.700 |
| | | 1.75 | 63 | 81 | 1.286 | 0.693 |
| | | 2.00 | 42 | 56 | 1.310 | 0.689 |
| 1 | 0.4 | 1.25 | 369 | 560 | 1.518 | 0.610 |
| | | 1.50 | 115 | 179 | 1.557 | 0.620 |
| | | 1.75 | 63 | 98 | 1.556 | 0.595 |
| | | 2.00 | 42 | 68 | 1.619 | 0.600 |
| 1 | 0.6 | 1.25 | 369 | 706 | 1.913 | 0.507 |
| | | 1.50 | 115 | 231 | 2.009 | 0.504 |
| | | 1.75 | 63 | 128 | 2.032 | 0.513 |
| | | 2.00 | 42 | 86 | 2.048 | 0.514 |
| 1 | 0.8 | 1.25 | 369 | 916 | 2.482 | 0.423 |
| | | 1.50 | 115 | 299 | 2.600 | 0.409 |
| | | 1.75 | 63 | 168 | 2.667 | 0.418 |
| | | 2.00 | 42 | 119 | 2.833 | 0.407 |
| 1 | 1.0 | 1.25 | 369 | 1278 | 3.463 | 0.324 |
| | | 1.50 | 115 | 401 | 3.487 | 0.313 |
| | | 1.75 | 63 | 225 | 3.514 | 0.313 |
| | | 2.00 | 42 | 156 | 3.714 | 0.300 |
| 2 | 0.2 | 1.25 | 271 | 329 | 1.214 | 0.723 |
| | | 1.50 | 82 | 103 | 1.256 | 0.716 |
| | | 1.75 | 45 | 57 | 1.267 | 0.711 |
| | | 2.00 | 31 | 41 | 1.323 | 0.708 |
| 2 | 0.4 | 1.25 | 271 | 409 | 1.509 | 0.634 |
| | | 1.50 | 82 | 131 | 1.598 | 0.628 |
| | | 1.75 | 45 | 72 | 1.600 | 0.621 |
| | | 2.00 | 31 | 48 | 1.548 | 0.613 |
| 2 | 0.6 | 1.25 | 271 | 516 | 1.904 | 0.528 |
| | | 1.50 | 82 | 168 | 2.049 | 0.521 |
| | | 1.75 | 45 | 92 | 2.044 | 0.495 |
| | | 2.00 | 31 | 64 | 2.065 | 0.503 |
| 2 | 0.8 | 1.25 | 271 | 684 | 2.524 | 0.434 |
| | | 1.50 | 82 | 223 | 2.720 | 0.418 |
| | | 1.75 | 45 | 124 | 2.756 | 0.421 |
| | | 2.00 | 31 | 83 | 2.677 | 0.429 |

| | | | | | | |
|---|---|---|---|---|---|---|
| 2 | 1.0 | 1.25 | 271 | 918 | 3.387 | 0.329 |
| | | 1.50 | 82 | 306 | 3.732 | 0.312 |
| | | 1.75 | 45 | 168 | 3.733 | 0.340 |
| | | 2.00 | 31 | 116 | 3.742 | 0.329 |
